# Elevated plasma phosphorylated tau 181 in amyotrophic lateral sclerosis relates to lower motor neuron dysfunction

**DOI:** 10.1101/2022.04.10.22273671

**Authors:** Katheryn A.Q. Cousins, Leslie M. Shaw, Sanjana Shellikeri, Laynie Dratch, Luis Rosario, Lauren B. Elman, Colin Quinn, Defne A. Amado, David A. Wolk, Thomas F. Tropea, Alice Chen-Plotkin, David J. Irwin, Murray Grossman, Edward B. Lee, John Q. Trojanowski, Corey T. McMillan

**Affiliations:** Department of Neurology, Perelman School of Medicine, University of Pennsylvania; Department of Pathology and Laboratory Medicine, University of Pennsylvania

## Abstract

**Objective:** Plasma phosphorylated tau (p-tau_181_) is reliably elevated in Alzheimer’s disease (AD), but less explored is its specificity relative to other neurodegenerative conditions. Here we find novel evidence that plasma p-tau_181_ is elevated in amytrophic lateral sclerosis (ALS), a neurodegenerative condition typically lacking tau pathology. We performed a detailed clinical evaluation to unravel the potential source of this unexpected observation.

**Methods:** Patients were clinically or pathologically diagnosed with ALS (n=130) or AD (n=82), or were healthy non-impaired controls (n=33). Receiver operating characteristic (ROC) curves were analyzed and area under the curve (AUC) was used to discriminate AD from ALS. Within ALS, Mann-Whitney-Wilcoxon tests compared analytes by presence/absence of upper motor neuron (UMN) and lower motor neuron (LMN) signs. Spearman correlations tested associations between plasma p-tau_181_ and postmortem neuron loss.

**Results:** A Wilcoxon test showed plasma p-tau_181_ was higher in ALS than controls (W=3297, *p*=0.0000020), and ROC analyses showed plasma p-tau_181_ poorly discriminated AD and ALS (AUC=0.60). In ALS, elevated plasma p-tau_181_ was associated with LMN signs in cervical (W=827, *p*=0.0072), thoracic (W=469, *p*=0.00025), and lumbosacral regions (W=851, *p*=0.0000029). In support of LMN findings, plasma p-tau_181_ was associated with neuron loss in the spinal cord (rho=0.46, *p*=0.017), but not in the motor cortex (*p*=0.41). CSF p-tau_181_ and plasma neurofilament light chain (NfL) were included as reference analytes, and demonstrate specificity of findings.

**Interpretation:** We found strong evidence that plasma p-tau_181_ is elevated in ALS and may be a novel marker specific to LMN dysfunction.

## 1. Introduction

Emerging plasma-based biomarkers of neurodegenerative disease meet the need for inexpensive, less invasive, and accurate diagnostic tools. In particular, elevated phosphorylated tau at threonine 181 (p-tau_181_) in plasma has been shown to be a highly accurate marker of Alzheimer’s disease (AD),^1,2^ and increased plasma p-tau_181_ is associated with amyloid-β plaques and tau-positive tangle neuropathology.^3^ Likewise, plasma p-tau_181_ is highly correlated with cerebrospinal spinal fluid (CSF) p-tau_181_ concentrations in AD,^4^ further supporting its reliability as a marker of AD brain alterations. However, unlike CSF which has direct contact with the brain, analyte concentrations in plasma may be influenced by disease events peripheral to the brain, not yet described.

A recent study from our group tested diagnostic accuracy of plasma p-tau_181_ in AD, but unexpectedly found high levels of p-tau_181_ in 11 patients with amyotrophic lateral sclerosis (ALS),^5^ a neurodegenerative condition involving upper motor neuron (UMN) and lower motor neuron (LMN) disease.^6^ Here we expand on this preliminary observation; we show new evidence that plasma p-tau_181_ is elevated in ALS patients. This is despite the fact that ALS is associated with TAR DNA-binding protein 43 (TDP-43) or SOD1 pathology, with little tau accumulation.^7^

We are aware of no previous studies that have investigated plasma p-tau_181_ in ALS, however our finding of elevated plasma p-tau_181_ is in contrast to consistently low concentrations of CSF p-tau_181_ observed in ALS.^8,9^ Moreover, studies of plasma p-tau_181_ in frontotemporal lobar degeneration associated with TDP-43 (FTLD-TDP) show that plasma p-tau_181_ is significantly lower in FTLD-TDP than AD,^10^ mirroring the relationship seen in CSF.^11^ These findings may indicate a potential disassociation between CSF and plasma p-tau181 in ALS, but not AD or FTLD.

In the first part of this study, we compared analyte concentrations and their diagnostic accuracy in ALS, AD, and cognitively unimpaired, healthy controls. Analyses compared plasma p-tau_181_ to plasma NfL, which has been previously identified as a marker in ALS,^12–14^ and to CSF p-tau_181_. We confirmed that elevated plasma p-tau_181_ was not due to co-occurring AD pathology by repeating comparisons in a subset of patients with autopsy-confirmed ALS and AD.

Given our finding that plasma p-tau_181_ was highly elevated in ALS, the second part of this study focused on ALS. We performed a detailed analysis to explore if plasma p-tau_181_ was related to clinical or pathologic features of ALS. First, correlations investigated divergences between plasma and CSF biomarkers within ALS. We next tested whether plasma p-tau_181_ was associated with lower motor neuron (LMN) and/or upper motor neuron (UMN) dysfunction in ALS. To test the specificity of our findings, we compared plasma p-tau_181_ to plasma NfL and CSF p-tau_181_ in ALS. We next investigated the clinical significance of plasma p-tau_181_ by testing its association with ALS clinical outcomes, including seated forced vital capacity (FVC) and the revised ALS functional rating scale (ALSFRS-R). Finally, in the subset of ALS patients with autopsy data, we tested if concentrations of plasma p-tau_181_ related to neuron loss in the motor cortex or spinal cord.

## 2. Materials and methods

### 2.1 Participants

Participants were 130 ALS patients who were seen at the the University of Pennsylvania ALS Center, Frontotemporal Degeneration Center, or AD Research Center; 82 AD patients and 33 non-impaired controls were included as reference groups. Inclusion criteria were a clinical diagnosis of ALS,^6,15^ AD,^16^ or normal cognition and plasma assayed for p-tau_181_. Informed consent was obtained according to the Declaration of Helsinki and approved by the Penn Institutional Review Board. Targeted genetic screening for known ALS and AD mutations identified 32 *C9orf72* repeat expansions and 5 *SOD1* point mutations in ALS patients, and 2 *PSEN1* mutations in AD patients.

Exclusion criterion for ALS patients was evidence of AD pathology, determined either by autopsy or by CSF amyloid-β 1-42 (Aβ_42_)≤192;^17^ 60 ALS patients were clinically-defined only, with neither autopsy or CSF data. The exclusion criterion for AD patients was negative evidence of AD pathology, determined either by autopsy or by CSF Aβ_42_>192. Exclusion criteria for controls was positive evidence of AD pathology, determined by CSF Aβ_42_≤192, or evidence of cognitive impairment, determined by mini-mental state exam (MMSE)<27.^18^

### 2.2 Neuropathological Assessment

A subset of patients had autopsy-confirmed pathology (28 ALS; 25 AD). Neuropathological diagnosis of ALS was according to criteria.^15,19^ Modern neuropathologic criteria for AD neuropathologic change (ADNC) were used to determine AD pathology;^20^ AD patients had high/intermediate ADNC and ALS patients had not/low ADNC. Braak stage (0-VI) assessed extent of tau spread in both AD and ALS.^21^ AD patients were Braak Stage V/VI (n=23); ALS patients were Braak Stage 0 (n=15), I/II (n=9), or III/IV (n=4). Neuronal loss was assessed in the motor cortex and anterior horn of the spinal cord in 26 ALS patients.^22^

### 2.3 Plasma and CSF collection and assay

For all patients, p-tau_181_ plasma concentrations were quantified using the single-molecule array (Simoa) platform, according to Advantage V2 Reagent Kits from Quanterix. Plasma NfL was also quantified using the Simoa Quanterix platform in a subset of ALS (n=120) and AD patients (n=70).

A subset of patients had CSF NfL data available, quantified using the Simoa Quanterix platform (12 ALS; 36 AD). CSF Aβ_42_ (50 ALS; 71 AD), p-tau_181_ (47 ALS; 70 AD), and total tau (t-tau) (48 ALS; 69 AD) were quantified using the xMAP Luminex platform.^23^ There was a mean of 13 months (SD=22.9) between plasma and CSF samples. A subset of these data is previously described.^5^

Technicians who performed biofluid analyses were blinded to clinical and pathological data.

### 2.4 Clinical Data

Demographic characteristics were age at symptom onset (defined by the first reported symptom), age at plasma collection (years), duration at plasma (years from symptom onset to plasma collection), and survival (years from symptom onset to death). One patient was missing disease onset year; 11 were missing onset date.

Clinical characteristics were available for subsets of ALS patients, assessed at clinical visit closest to plasma collection (mean=0.8 months; SD=3.9): seated forced vital capacity (FVC; n=109),^24^ revised ALS functional rating scale (ALSFRS-R; max score=48; n=118),^25^ ALSFRS-R Progression Rate (48 − ALSFRS-R/duration at ALSFRS-R in months; n=115), change in weight from baseline (weight at visit − weight at initial visit; n=80), and Penn UMN Total Score closest to visit (max=32; n=114).^26^ A neuromuscular examination by a board-certified neurologist tested for the presence or absence of UMN and LMN signs related to bulbar, cervical, thoracic, and lumbosacral regions (n=117); UMN thoracic was not assessed.^27^ There was an average of 0.8 months (standard deviation [SD]=3.89) between plasma collection and ALS clinical visits.

Initial clinical diagnoses for ALS patients were recorded as ALS (n=82), progressive muscular atrophy (PMA; n=13),^28^ primary lateral sclerosis (PLS; n=11),^29^ frontotemporal dementia (FTD; n=6),^30^ and progressive bulbar palsy (PBP; n=6).^31^

### 2.5 Statistical Analyses

Non-parametric tests were used to account for non-normal distribution of demographic, clinical, and biofluid variables. All statistical tests were performed with a significance threshold of α=0.05. Mann–Whitney–Wilcoxon tests performed comparisons between ALS and AD for continuous demographic variables and Chi-square tests compared categorical variables. For pairwise comparisons, effect sizes were calculated using rank-biserial correlation (*r*_*rb*_) (>0.1 was considered small; >0.2 medium; >0.3 large)^32^. Missing data were dropped from analyses. Analyses were conducted using R version 4.1.2 (2021-11-01) statistical software and the cutpointr,^33^ effectsize,^34^ and survival packages.^35^

#### 2.5.1 Analyte comparisons across ALS and AD; ALS and Controls

Part 1 of this study tested for elevated plasma p-tau_181_ in ALS. Wilcoxon tests performed planned pairwise comparisons of ALS to AD, and to Controls; CSF p-tau∼181, plasma NfL and CSF NfL were included as reference analytes. We confirmed findings in the subset of autopsy-confirmed ALS patients with Braak Stage ≤ II and autopsy-confirmed AD. Next, receiver operating characteristic (ROC) analyses tested diagnostic performance of p-tau_181_ and NfL in both plasma and CSF when discriminating AD patients from ALS patients. ROC metrics were calculated using bootstrapping with 250 iterations: area under the curve (AUC), AUC 95% confidence interval (CI), and best threshold determined by Youden’s index to maximize sensitivity and specificity.

#### 2.5.2 Analyses of plasma p-tau_181_ within ALS

In part 2 of this study, we focused analyses within ALS patients to explore the clinical and pathological correlates of plasma p-tau_181_.

##### Correlations between plasma and CSF

We tested for a divergence between plasma p-tau_181_ and CSF analytes in ALS using Spearman’s rho correlations. We compared this to correlations between plasma NfL and CSF analytes.

##### Associations with LMN and UMN signs

Wilcoxon tests investigated whether p-tau_181_ levels were associated with the presence/absence of LMN and/or UMN signs in ALS. To confirm findings, we also compared plasma p-tau_181_ levels across onset site and across initial diagnoses in patients with an ultimate diagnosis of ALS. To test specificity of plasma p-tau_181_ we included plasma NfL and CSF p-tau_181_ as reference analytes.

Co-occurring tau pathology has sometimes been observed in ALS patients with cognitive impairment.^36–38^ We therefore tested plasma p-tau_181_ in patients with clinical diagnoses of ALS with mild cognitive impairment (ALS-MCI) or ALS with FTD (ALS-FTD) (n=37) compared to ALS patients without evidence of cognitive impairment (n=93).

##### Clinical correlates of plasma p-tau_181_

We tested how plasma p-tau_181_ predicted clinical outcomes in ALS. Linear models then tested how plasma p-tau_181_ related to each clinical outcome (ALSFRS-R, ALSFRS-R Progression Rate, change in weight, FVC, UMN Total). All linear models included the interval from plasma collection to clinical visit (years), disease duration (months), and sex (male, female) as covariates. To perform non-parametric linear models, all continuous variables were rank-transformed.^39^

##### Pathological correlates of plasma p-tau_181_

In the subset of autopsy patients with histopathological data, Spearman correlations tested associations between plasma p-tau_181_ with neuron loss in the motor cortex and spinal cord. Finally to ensure that AD pathology did not explain variance in plasma levels observed in ALS, Kruskal-Wallis tests investigated if plasma p-tau_181_ differed by Thal Phase, CERAD score, or Braak Stage. All comparisons and linear models were also tested in plasma NfL and CSF p-tau.

## 3. Results

Table 1 summarizes demographic, CSF, and pathologic characteristics of ALS, AD and Controls. Of note, the majority of AD patients in our sample were early-onset (n=67; 82%),

**Table 1:**
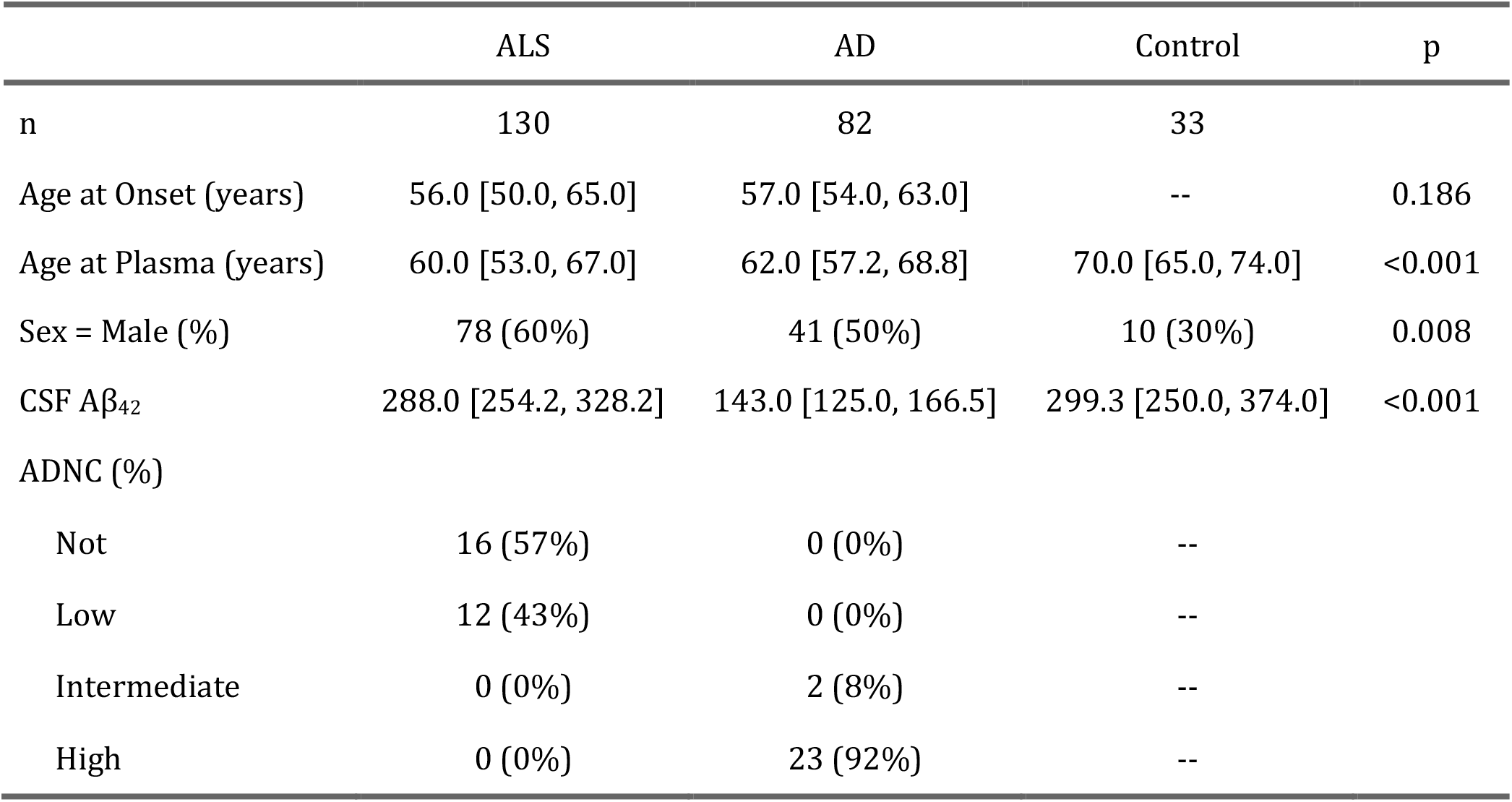
Characteristics of AD and ALS patients. Demographic, CSF, and pathological characteristics of patients. Missing data were dropped. For continuous variables, median and interquartile range (median [IQR]) are provided; *p*-values are reported for Mann-Whitney-Wilcoxon tests. For categorical variables, count and percentage (%) are provided; *p*-values are reported from chi-square tests.

### 3.1 Plasma p-tau_181_ in ALS compared with AD and non-impaired Controls

Figure 1 shows comparisons for biomarkers across ALS, AD and controls. Wilcoxon tests showed that plasma p-tau_181_ was higher in ALS than controls (W=3297, *p*=0.0000020) with large effect size (*r*_*rb*_=0.54); in contrast, CSF p-tau_181_ was lower in ALS than controls (W=250.5, *p*=0.00000028) with large effect size (*r*_*rb*_=0.54). Plasma p-tau_181_ was higher in AD than ALS (W=4305.5, *p*=0.019) with small effect size (*r*_*rb*_=0.19), while CSF p-tau_181_ was significantly higher in AD than ALS (W=71.5, *p*=2.1e-18) with large effect size (*r*_*rb*_=0.96). As previously reported, NfL was higher in ALS than AD in both plasma (W=7395.5, *p*=2.4e-18; *r*_*rb*_=0.76) and CSF (W=350, *p*=0.00095; *r*_*rb*_=0.62).

**Figure 1:**
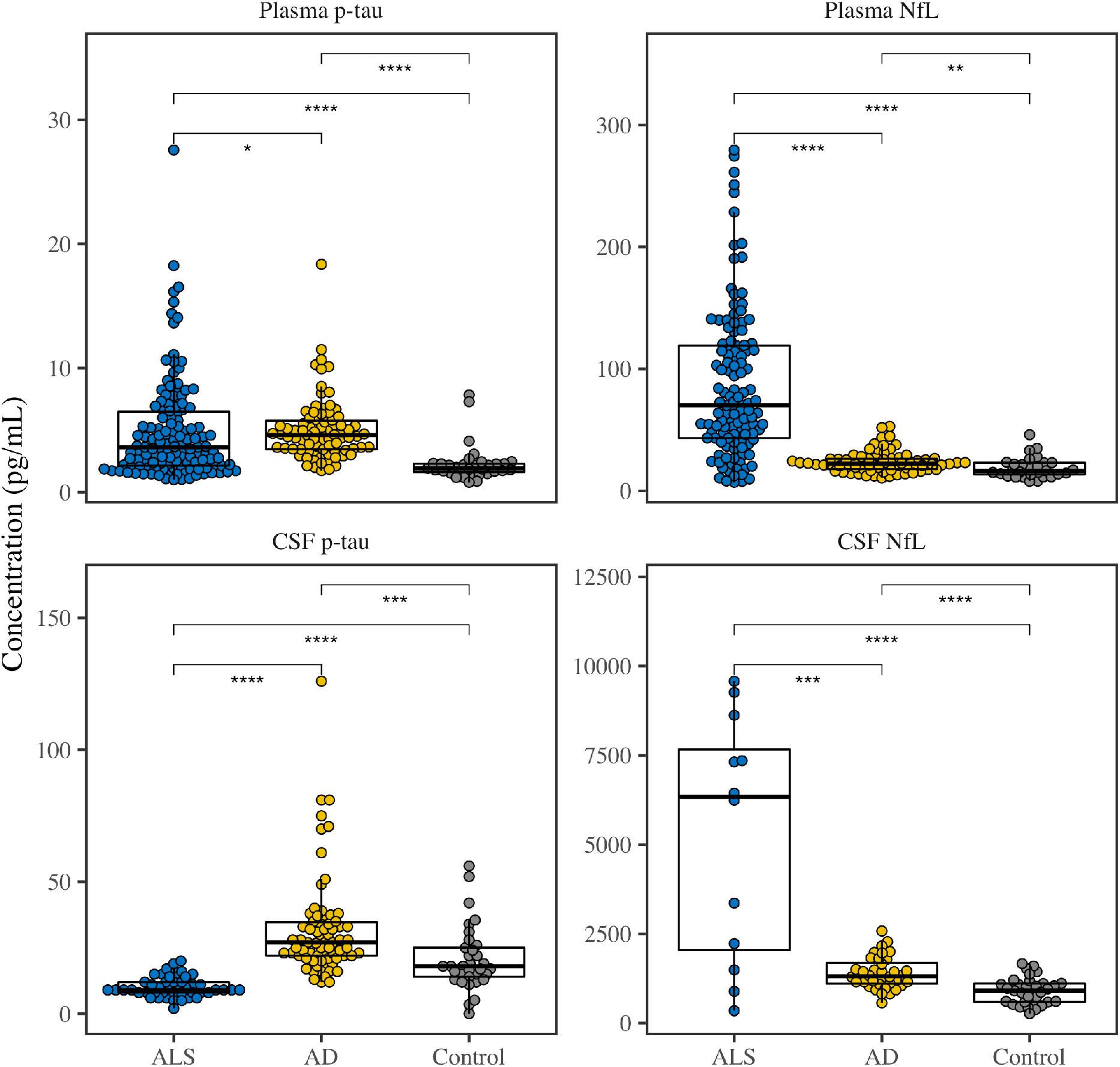
Plasma and CSF p-tau_181_ and NfL levels. Comparisons of plasma concentrations (top panels) and CSF concentrations (bottom panels) of p-tau_181_ (left panels) and NfL (right panels) across ALS (blue), AD (yellow), and Controls (grey). Asterisks represent *p*-values from Wilcoxon pairwise comparisons (\**p*<0.05, \*\**p*<0.01, \*\*\**p*<0.001, \*\*\*\**p*<0.0001). Boxplots show median and interquartile range (IQR).

To ensure elevated plasma p-tau_181_ in ALS was not due to concomitant AD, we repeated analyses in the subset of autopsy-confirmed AD patients (n=25) and in autopsy-confirmed ALS patients with minimal tau (Braak Stage 0/I/II) and minimal AD (not/low ADNC) (n=24). A Wilcoxon test confirmed that plasma p-tau_181_ levels were no different (W=279, *p*=0.68) between ALS (median=6.4; IQR=5.28) and AD (median=5; IQR=3).

Next, ROC analyses compared CSF and plasma metrics when discriminating ALS and AD. Plasma p-tau_181_ had the lowest diagnostic accuracy (AUC=0.60), highlighting that plasma p-tau_181_ is elevated in both ALS and AD. Unlike plasma p-tau_181_, CSF p-tau_181_ had excellent diagnostic discrimination between AD and ALS (AUC=0.98), as did plasma NfL (AUC=0.88).

### 3.2 Correlates of plasma p-tau_181_ within ALS

In part 2 of this study, we focused analyses within ALS patients to explore the clinical and pathological correlates of plasma p-tau_181_.

#### 3.2.1 Associations of plasma with CSF in ALS

To better understand the dissociation between plasma and CSF p-tau_181_ in ALS, we tested correlations between plasma p-tau_181_ and NfL with CSF analytes. Spearman’s rho tests showed that plasma p-tau_181_ was not correlated with any CSF metrics: CSF Aβ_42_ (*p*=0.70), CSF p-tau_181_ (*p*=0.90), CSF t-tau (*p*=0.59), or CSF NfL (*p*=0.99). Plasma p-tau_181_ was also not correlated with plasma NfL in ALS (*p*=0.32). In contrast to the null results for plasma p-tau_181_, plasma NfL was positively correlated with both CSF t-tau (rho=0.37, *p*=0.0088) and CSF NfL (rho=0.69, *p*=0.016); plasma NfL was not significantly correlated with CSF Aβ_42_ (*p*=0.72) or CSF p-tau_181_ in ALS (*p*=0.82).

#### 3.2.2 Lower Motor Neuron (LMN) dysfunction in ALS

We next tested if plasma p-tau_181_ levels differed by the presence or absence of LMN and UMN signs in ALS (Figure 2A). To test specificity of plasma p-tau_181_ findings, we also tested plasma NfL (Figure 2B) and CSF p-tau_181_ (Figure 2C).

**Figure 2:**
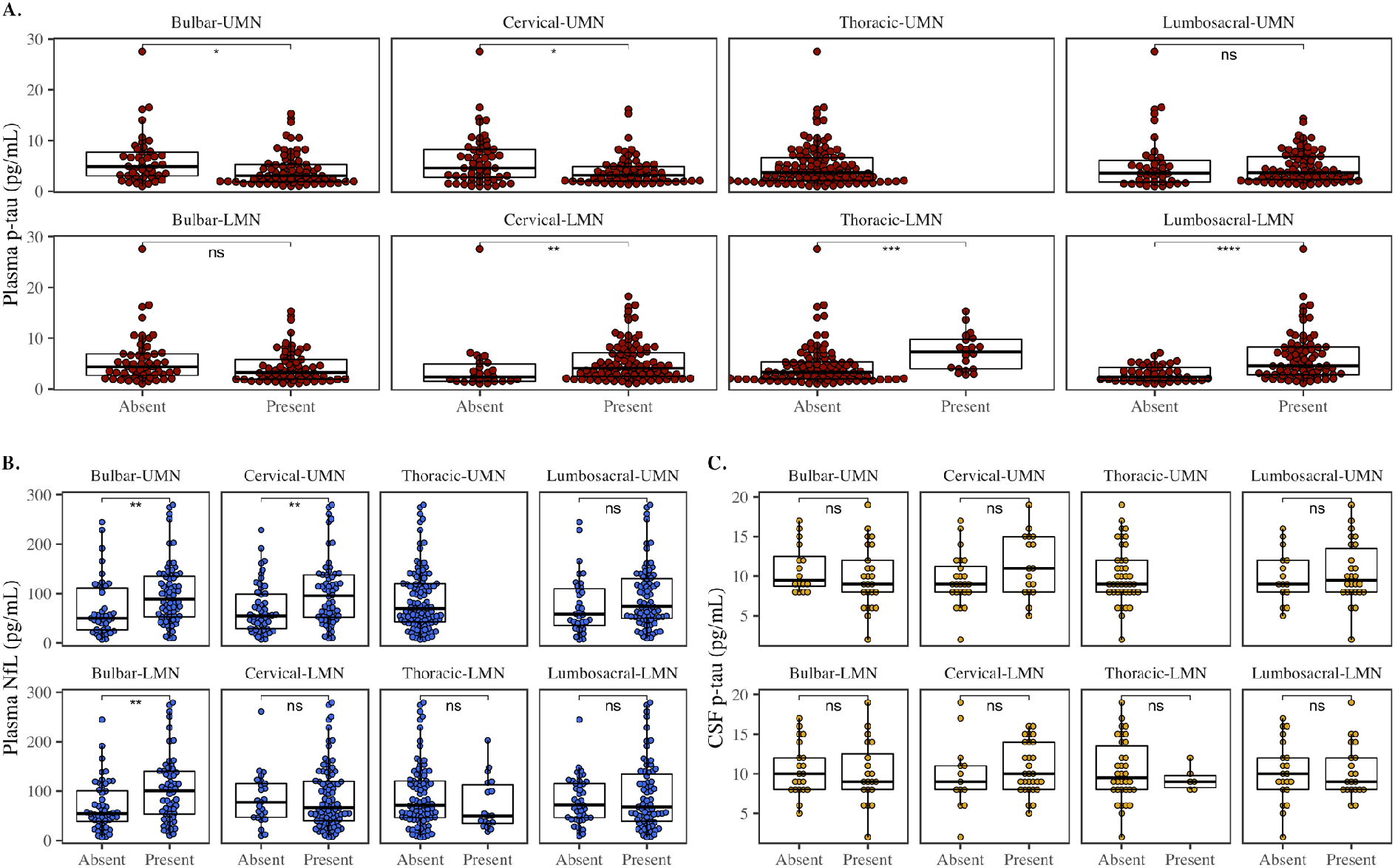
Plasma and CSF levels by Motor Examination. Comparisons of concentrations across UMN and LMN regions: (A.) p-tau_181_ in red, (B.) plasma NfL in blue, and (C.) CSF p-tau_181_ in yellow. Boxplots show median and interquartile range (IQR). Asterisks represent p-values from Wilcoxon pairwise comparisons (\**p*<0.05, \*\**p*<0.01, \*\*\**p*<0.001, \*\*\*\**p*<0.0001, or not significant [ns]).

There were significantly elevated plasma p-tau_181_ levels in ALS patients who showed motor examination findings of LMN signs related to cervical (W=827, *p*=0.0072; *r*_*rb*_=0.34), thoracic (W=469, *p*=0.00025; *r*_*rb*_=0.52), and lumbosacral regions (W=851, *p*=0.0000029; *r*_*rb*_=0.51); there was no difference based on presence or absence of bulbar LMN signs (*p*=0.089). Plasma p-tau_181_ was significantly lower for patients who were positive for UMN signs in bulbar (W=2126, *p*=0.012; *r*_*rb*_=0.28) and cervical regions (W=2258, *p*=0.023; *r*_*rb*_=0.24); there was no difference for UMN lumbosacral signs (*p*=0.71).

In contrast to plasma p-tau_181_, plasma NfL levels (Figure 2B) did not differ based on LMN signs in cervical (*p*=0.87), thoracic (*p*=0.27), or lumbosacral regions (*p*=0.86). Instead, NfL was elevated in patients positive for bulbar signs, both LMN (W=1019, *p*=0.0034; *r*_*rb*_=0.32) and UMN (W=935, *p*=0.0041; *r*_*rb*_=0.33), as well as cervical UMN signs (W=1062, *p*=0.0057; *r*_*rb*_=0.31); there was no difference based on lumbosacral UMN signs (*p*=0.15). CSF p-tau_181_ levels (Figure 2C) did not differ by UMN or LMN signs (all *p*>0.2).

Results suggested that plasma p-tau_181_ related to LMN limb/thorax dysfunction, a finding distinct from plasma NfL which related to both UMN and LMN bulbar signs. To test the consistency of this finding, we tested how plasma p-tau levels varied by initial diagnosis and onset sites. If plasma p-tau_181_ is associated with LMN signs, then we expected higher levels of plasma p-tau_181_ in ALS patients with a LMN onset or an initial diagnosis of PMA, compared to initial PLS or initial cognitive signs (FTD or PBA).

Patients with LMN limb onset in cervical and lumbosacral regions had higher median plasma p-tau_181_ values than UMN onset (Figure 3A); only 1 patient had thoracic LMN onset, but this patient had relatively high plasma p-tau_181_. This difference in plasma p-tau_181_ by LMN *vs*. UMN onset was confirmed by a Wilcoxon Test (W=271, *p*=0.00052) with large effect size (*r*_*rb*_=0.52). Neither plasma NfL (W=373, *p*=0.097) nor CSF p-tau_181_ (*p*=0.86) differed by LMN *vs*. UMN onset. However, plasma NfL medians were highest in patients with both UMN and LMN bulbar onset (Figure 3B); one patient with LMN thoracic onset also had high NfL levels.

**Figure 3:**
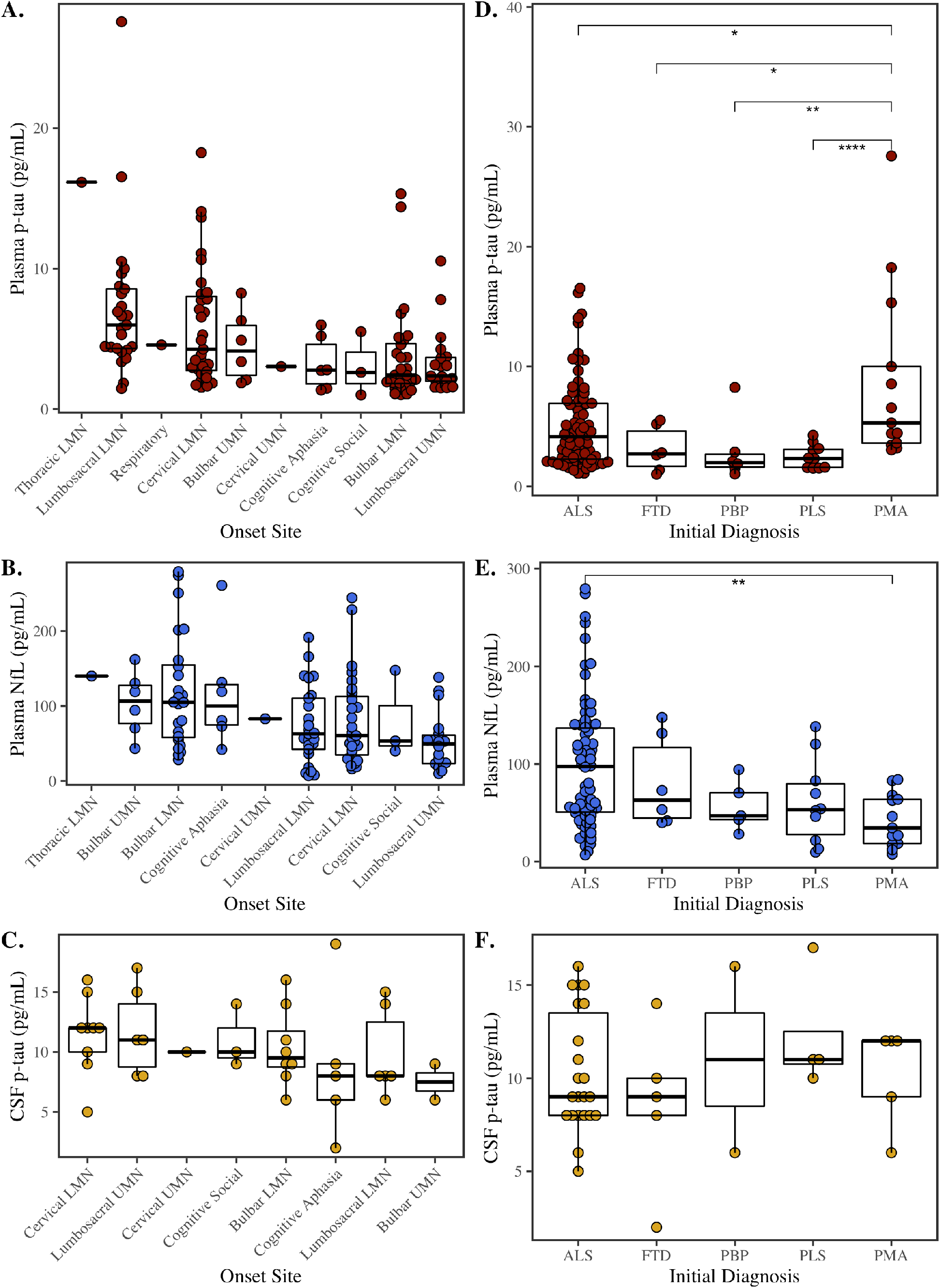
Differences in plasma and CSF by initial diagnosis and by onset site. Comparisons across onset site, ordered by highest-to-lowest median: (A.) plasma p-tau_181_ in red, (B.) plasma NfL in blue, and (C.) CSF p-tau_181_ in yellow. Comparisons across initial diagnosis in patients who convert to ALS: (D.) plasma p-tau_181_ in red, (E.) plasma NfL in blue, and (F.) CSF p-tau_181_ in yellow. Asterisks represent p-values from Wilcoxon pairwise comparisons (\**p*<0.05, \*\**p*<0.01, \*\*\**p*<0.001, \*\*\*\**p*<0.0001, or not significant [ns]). Boxplots show median, interquartile range (IQR), and outliers.

Plasma p-tau_181_ differed by initial diagnosis (Figure 3D), confirmed by a Kruskal-Wallis test (χ^2^(4)=18, *p*=0.0011). Planned pairwise comparisons showed that ALS patients with initial PMA, characterized by LMN-only onset, had significantly higher p-tau_181_ levels than all other initial diagnoses, including PLS (W=9, *p*=0.000078; *r*_*rb*_=0.87), PBP (W=8, *p*=0.0047; *r*_*rb*_=0.79), FTD (W=13, *p*=0.022; *r*_*rb*_=0.67), and ALS (W=348.5, *p*=0.046; *r*_*rb*_=0.35). Plasma NfL also differed by initial diagnosis (Figure 3E; χ^2^(4)=14, *p*=0.0072), where an initial diagnosis of ALS was associated with the higher levels of plasma NfL than PMA (W=750, *p*=0.0014; *r*_*rb*_=0.56); no other groups were significantly different. CSF p-tau_181_ did not vary by initial diagnosis (Fig 3F; *p*=0.65).

Finally, we tested if analyte levels differed in ALS patients with cognitive impairment (clinical diagnosis of ALS-FTD or ALS-MCI) compared to ALS patients without cognitive impairment.^36–38^ A Wilcoxon test showed that plasma p-tau_181_ levels were lower in ALS-MCI/ALS-FTD (median=2.8; IQR=2.54) compared to ALS without cognitive impairment (median=3.72; IQR=5.09) (W=2119, *p*=0.04), consistent with no association with UMN. For plasma NfL, there was no significant difference between ALS-MCI/ALS-FTD (median=80.6; IQR=55.5) and ALS (median=57.5; IQR=82.2) (W=1235, *p*=0.088); nor for CSF p-tau_181_ (*p*=0.39).

#### 3.2.3 Clinical correlates of plasma p-tau_181_

We examined how demographic variables in ALS influenced plasma p-tau_181_ within ALS. Plasma p-tau_181_ was higher in men than women (W=1421.5, *p*=0.0040); the association with disease duration was not significant (rho=0.17, *p*=0.052). Plasma p-tau_181_ was not correlated with age at plasma collection (*p*=0.11). Individuals with an *SOD1* point mutation, typically characterized by greater LMN dysfunction,^40^ had significantly higher plasma p-tau_181_ levels (median=6.93; IQR=1.85) than apparent sporadic individuals (median=3.44; IQR=4.22) (W=103.5, *p*=0.046); there was no difference between individuals with a *C9orf72* repeat expansion, typically characterized by greater UMN dysfunction, and apparent sporadic ALS individuals (*p*=0.38).

We tested how plasma p-tau_181_ correlated with clinical outcomes, with uncorrected Spearman correlations reported (Figure 4). Linear models tested each clinical outcome as a function of p-tau_181_, covarying for the interval from plasma to clinical collection, disease duration at clinical visit, and sex. More severe weight loss was significantly associated with increased plasma p-tau_181_ (β=-0.24, SE=0.074, *p*=0.0019); no other covariates were significant (all *p*>0.2). Worse ALSFRS-R was significantly associated with increased plasma p-tau_181_ (β=-0.17, SE=0.081, *p*=0.044), covarying for plasma-to-visit interval (*p*=0.66), disease duration (β=-0.37, SE=0.087, *p*=0.000035), and sex (β=14, SE=6, *p*=0.024). While the Spearman’s correlation was not significant (see Figure 4), ALSFRS-R Progression Rate was significantly associated with plasma p-tau_181_ (β=0.16, SE=0.071, *p*=0.025), after covarying for plasma-to-visit interval (*p*=0.84), disease duration (β=-0.6, SE=0.076, *p*=2.5e-12), and sex (β=-14, SE=5.3, *p*=0.012). Reduced FVC was significantly associated with increased plasma p-tau_181_ (β=-0.25, SE=0.086, *p*=0.0045); no other covariates were associated with FVC (all *p*>0.3). UMN total score was inversely associated with plasma p-tau_181_ (β=-0.22, SE=0.083, *p*=0.0095) and was lower in men than women (β=-17, SE=6.2, *p*=0.0084); no other covariates were associated with UMN total (all *p*>0.3).

**Figure 4:**
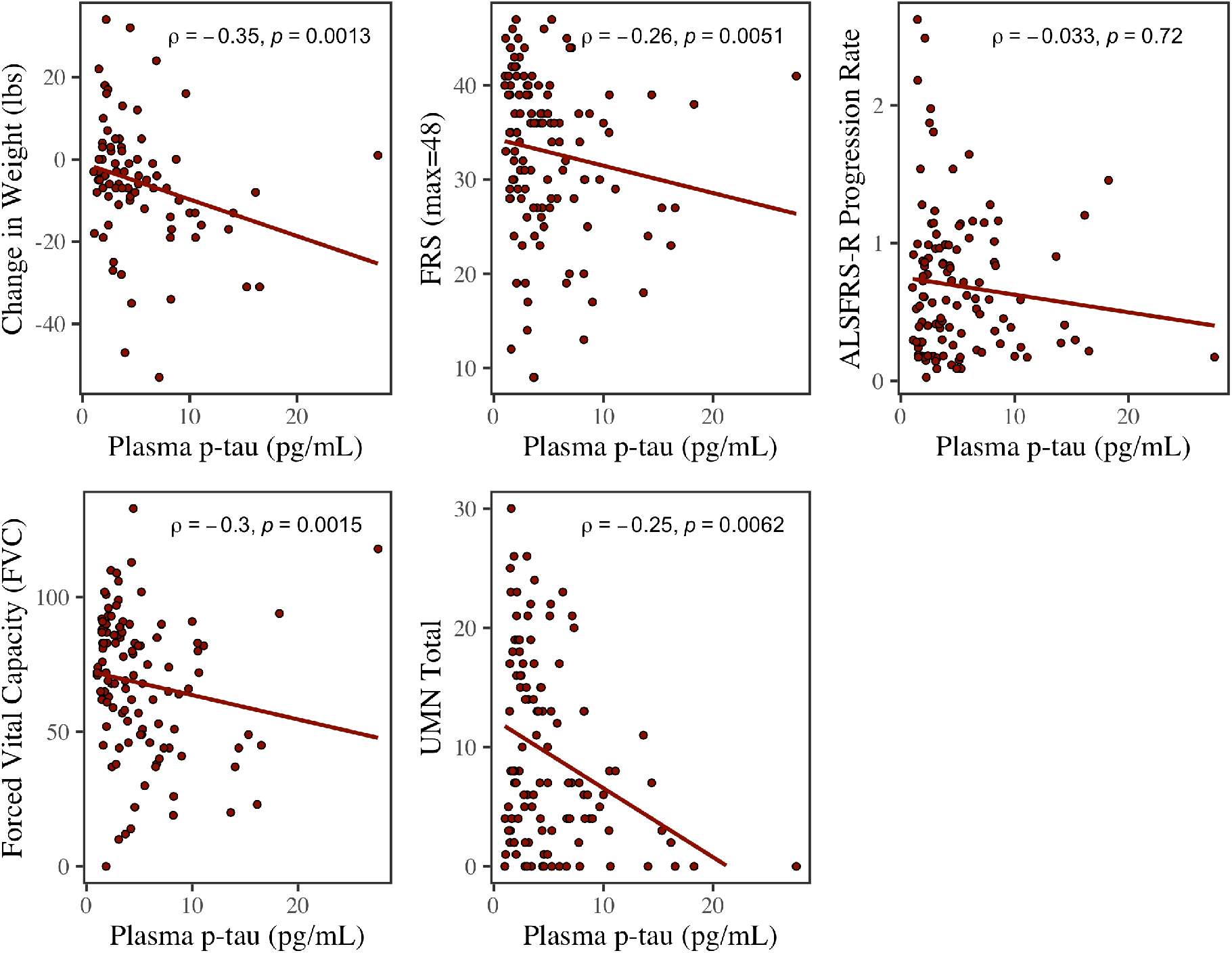
Plasma levels by clinical outcomes. Concentrations of plasma p-tau_181_ across clinical outcomes: Change in Weight, ALSFRS-R, ALSFRS-R Progression Rate, Forced Vital Capacity (FVC), and Upper Motor Neuron (UMN) Total. Least squares regression lines are plotted in red; Spearman rho (ρ) correlations are reported.

#### 3.2.4 Pathological correlates of plasma p-tau_181_ in autopsy-confirmed ALS

Spearman correlations indicated that increased p-tau_181_ was associated with greater neuron loss in the spinal cord (rho=0.46, *p*=0.017), but was not significantly associated with motor cortex neuron loss (*p*=0.41), further supporting the relationship with LMN disease (Figure 5).

**Figure 5:**
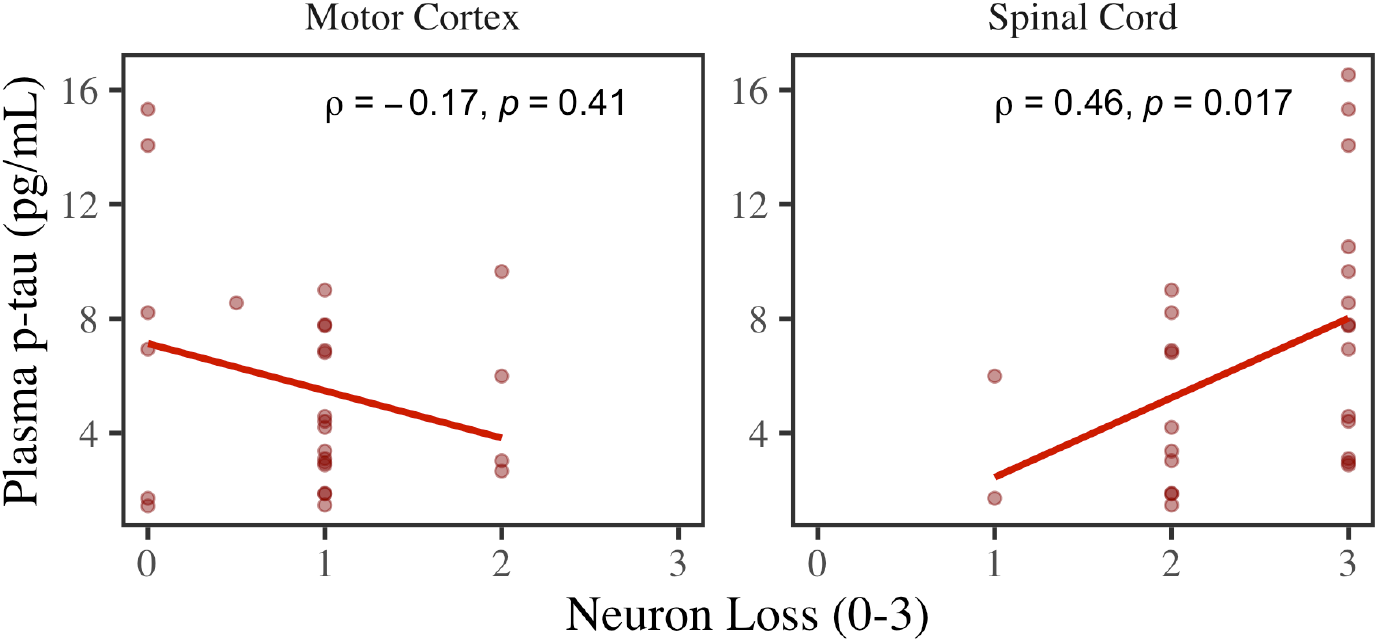
Associations with atrophy in motor cortex and spinal cord. Scatterplots of neuron loss (0-3) in motor cortex (left panel) and the anterior horn of the spinal cord (right panel) and their associations with plasma p-tau_181_. Least squares regression lines are plotted for each panel; Spearman rho (ρ) correlations are reported.

Finally, Kruskal-Wallis tests showed that plasma p-tau_181_ levels did not differ by Braak stage (*p*=0.57), CERAD score (*p*=0.25), or Thal phase (*p*=0.96), further confirming that elevated plasma p-tau_181_ in ALS was not due to low levels of ADNC in ALS patients.

## 4. Discussion

While much work indicates that high p-tau_181_ in plasma is an accurate marker of AD,^1,2^ our study presents novel evidence that plasma p-tau_181_ is also elevated in some ALS patients. Indeed, patients with ALS had significantly elevated plasma p-tau_181_ levels compared to non-impaired healthy controls. While AD had higher plasma p-tau_181_ than ALS, the effect size was small. Importantly, there was no difference in the subset of autopsy-confirmed AD and ALS, indicating that co-occurring AD was not driving elevated plasma p-tau_181_ in ALS. Moreover, ROC analyses showed that plasma p-tau_181_ had the poorest performance when discriminating ALS and AD patients (AUC=0.60). This is in contrast to CSF p-tau_181_, which was significantly higher in AD than ALS with large effect size and showed excellent discrimination (AUC=0.98). The divergence between plasma and CSF p-tau_181_ within ALS was further confirmed by correlations that showed no associations between plasma p-tau_181_ and any CSF biomarkers. In addition, we show strong and consistent evidence that plasma p-tau_181_ was related to limb/thorax LMN dysfunction and degeneration in ALS: plasma p-tau_181_ was highest in ALS patients with an initial diagnosis of PMA, with limb LMN onset, and with LMN signs in cervical, thoracic, and lumbosacral regions. It was also inversely correlated with UMN total score,^41^ and associated with greater neuronal loss in the spinal cord, but not with motor cortex. These associations with LMN disease were specific to plasma p-tau_181_, and CSF p-tau_181_ was not associated with signs in ALS in this study or others.^9^ Increased plasma p-tau_181_ was also associated with ALS clinical outcomes – including weight loss, reduced FVC, and lower ALSFRS-R – and in patients with a *SOD1* mutation. Taken together, our study makes two important contributions: 1.) that elevated plasma p-tau_181_ may not be specific to AD, and 2.) that elevated plasma p-tau_181_ may be a biomarker of LMN dysfunction and degeneration in ALS.

We saw evidence that plasma p-tau and plasma NfL are two distinct, but informative, markers in ALS. Unlike plasma p-tau_181_, plasma NfL was higher in ALS than AD, had good discrimination (AUC=0.88), and was significantly correlated with CSF NfL. Our findings are corroborated by other studies showing elevated plasma NfL^13^ and CSF NfL^42,43^ in ALS, and that plasma and CSF NfL are correlated.^44,45^ While the relationship between plasma p-tau_181_ and LMN dysfunction was robust and consistently observed across our analyses, previous work has shown that CSF and serum NfL is associated with UMN disease in ALS.^43,46^ Our data instead suggested that NfL was most consistently associated with both UMN and LMN bulbar signs, in addition to cervical UMN. Consistent with this finding, plasma NfL levels were highest in patients with bulbar onset. Thus, the relationship with LMN disease may be specific to plasma p-tau_181_, while plasma NfL may be most associated with bulbar onset and signs. Given the novelty of our findings, it will be important to validate in an independent sample, and future studies should test plasma p-tau_181_ as a marker of other motor neuron diseases or of axonal peripheral neuropathies. Importantly, the mechanism for elevated plasma p-tau_181_ in ALS must still be elucidated since ALS is typically associated with TDP-43 pathology.^47^ While there is some evidence of hyperphosphoylated tau pathology in ALS,^48,49^ this is typically observed in cases with cognitive impairment and FTD spectrum cases.^36–38^ We saw no evidence that higher plasma p-tau_181_ was linked to cognitive impairment in ALS, and we instead observed somewhat lower plasma p-tau levels in ALS-MCI/ALS-FTD patients. Our analyses show that higher plasma p-tau_181_ was associated with neuron loss in the spinal cord, however the sample size was small and there were only 28 cases with postmortem assessments. Future histopathological studies should test if pathological accumulation or neurodengeneration in the spinal cord relates to plasma p-tau_181_ levels in ALS. On the other end of the disease spectrum, it will be important to investigate plasma p-tau_181_ as it relates to early disease in ALS. Here we studied plasma p-tau_181_ in symptomatic ALS and AD, and ROC analyses showed poor discrimination by plasma p-tau_181_. While discrimination by plasma p-tau_181_ is less useful for symptomatic ALS and AD, which are clinically distinct entities, this overlap in plasma p-tau_181_ levels may be important to consider for prodromal/early-stages of disease. Prior work shows that plasma NfL was elevated in pre-symptomatic ALS patients who were less than 5 years from disease onset,^13^ and is sensitive to ALS disease processes.^14,50^ Future studies should test whether plasma p-tau_181_ is elevated in pre-symptomatic individuals with a familial history of ALS.

There are some caveats to our findings. First, we had no apriori hypothesis that plasma p-tau_181_ levels were elevated in ALS, and the mechanism for this effect is still unknown; findings should be validated in an independent sample. Second, the number of ALS patients with postmortem pathological data was small, and this limited our ability to investigate the association with spinal cord pathology that was specific to increased plasma p-tau_181_ in ALS. Future studies should test for a dissociation between plasma p-tau_181_ and plasma NfL related to pathology accumulation and degeneration in the spinal cord and LMN compared to the motor cortex. Despite only a subset of patients having postmortem pathological data, we repeated analyses in the subset of autopsy-confirmed ALS with low/not ADNC and found levels of plasma p-tau_181_ were similarly elevated compared to AD. This confirmed that high levels of plasma p-tau_181_ were not explained by co-occurring ADNC. Third, only a subset of our sample had CSF data. Even so, our data are supported by previous literature which finds that CSF p-tau_181_ is reduced in ALS compared to AD.^8,9^ Only 12 patients had both CSF NfL data and ALS clinical data, which precluded our ability to test for differences versus agreement between CSF NfL and plasma NfL related to LMN disease, UMN disease, and clinical outcomes in ALS. Fourth, we tested analyte levels by the presence or absence of LMN and UMN symptoms, however future studies should examine how plasma biomarkers in ALS relate to continuous metrics of UMN and LMN neurophysiology.^51^ Biomarkers of ALS are still needed to provide an early diagnosis, and to inform disease management and eligibility criteria for treatment trials. Moreover, because plasma is a minimally invasive biomarker available through blood draw, it is more easily collected repeatedly and in pre-symptomatic cases. Our study shows novel evidence that plasma p-tau_181_ is highly elevated in ALS and may be sensitive to LMN disease in ALS. Moreover, plasma p-tau_181_ as a marker of LMN disease may be complimentary to plasma NfL in ALS.

## Data Availability

All data produced in the present study are available upon reasonable request of qualified investigator to the authors.

## Acknowledgements/Conflicts/Funding Sources

This work is supported by funding from the National Institute of Aging (P01-AG066597, U19-AG062418, P30-AG072979, R01-AG054519, R01-AG052943) and the Penn Institute on Aging. KAQC is supported by the Alzheimer’s Association (AARF-D-619473, AARF-D-619473-RAPID) and P30-AG072979. TFT is supported by the NIH/NINDS (K23-NS11416-01A1).

## Author Contributions

KAQC, DJI, and CM had major contributions to the conception of the research question. KAQC, LMS, SS, DJI, MG, and CM all contributed to the methodology and design of the study. All authors, KAQC, LMS, SS, DJI, LD, LBE, CQ, DAA, LR, MG, DAW, TFT, AC, EBL, and CM, were involved in data acquisition. KAQC performed all formal statistical analyses and investigations in this study. KAQC prepared the original manuscript draft. All authors, KAQC, LMS, SS, DJI, LD, LBE, CQ, DAA, LR, MG, DAW, TFT, AC, EBL, and CM, were involved in manuscript review and editing and have approved the final draft.

## References

1. Brickman AM, Manly JJ, Honig LS, et al. Plasma p-tau181, p-tau217, and other blood-based Alzheimer’s disease biomarkers in a multi-ethnic, community study. Alzheimer’s and Dementia 2021;(September 2020):1–12.

2. Lantero Rodriguez J, Karikari TK, Suárez-Calvet M, et al. Plasma p-tau181 accurately predicts Alzheimer’s disease pathology at least 8 years prior to post-mortem and improves the clinical characterisation of cognitive decline [Internet]. Acta Neuropathologica 2020;140(3):267–278.Available from: https://doi.org/10.1007/s00401-020-02195-x

3. Thijssen EH, La Joie R, Strom A, et al. Plasma phosphorylated tau 217 and phosphorylated tau 181 as biomarkers in Alzheimer’s disease and frontotemporal lobar degeneration: a retrospective diagnostic performance study. The Lancet Neurology 2021;20(9):739–752.

4. Janelidze S, Mattsson N, Palmqvist S, et al. Plasma P-tau181 in Alzheimer’s disease: relationship to other biomarkers, differential diagnosis, neuropathology and longitudinal progression to Alzheimer’s dementia [Internet]. Nature Medicine 2020;26(3):379–386.Available from: http://dx.doi.org/10.1038/s41591-020-0755-1

5. Tropea TF, Waligorska T, Xie SX, et al. Plasma Phosphorylated Tau181 is a Biomarker of Alzheimer’s Disease Pathology and Associated with Cognitive and Functional Decline. 2022;

6. Strong MJ, Abrahams S, Goldstein LH, et al. Amyotrophic lateral sclerosis-frontotemporal spectrum disorder (ALS-FTSD): Revised diagnostic criteria [Internet]. Amyotrophic lateral sclerosis and frontotemporal degeneration 2017;18(3-4):153–174.Available from: https://www.tandfonline.com/doi/full/10.1080/21678421.2016.1267768

7. Liscic RM, Breljak D. Molecular basis of amyotrophic lateral sclerosis [Internet]. Progress in Neuro-Psychopharmacology and Biological Psychiatry 2011;35(2):370–372.Available from: https://www.sciencedirect.com/science/article/pii/S0278584610002800

8. Grossman M, Elman L, McCluskey L, et al. Phosphorylated tau as a candidate biomarker for amyotrophic lateral sclerosis. JAMA neurology 2014;71(4):442–448.

9. Scarafino A, D’Errico E, Introna A, et al. Diagnostic and prognostic power of CSF Tau in amyotrophic lateral sclerosis. Journal of neurology 2018;265(10):2353– 2362.

10. Thijssen EH, La Joie R, Wolf A, et al. Diagnostic value of plasma phosphorylated tau181 in Alzheimer’s disease and frontotemporal lobar degeneration. Nature Medicine 2020;26(3):387–397.

11. Cousins KAQ, Irwin DJ, Wolk DA, et al. ATN status in amnestic and non-amnestic Alzheimer’s disease and frontotemporal lobar degeneration. Brain 2020;143(7):2295–2311.

12. Lu C-H, Macdonald-Wallis C, Gray E, et al. Neurofilament light chain: a prognostic biomarker in amyotrophic lateral sclerosis. Neurology 2015;84(22):2247– 2257.

13. Bjornevik K, O’Reilly EJ, Molsberry S, et al. Prediagnostic Neurofilament Light Chain Levels in Amyotrophic Lateral Sclerosis. Neurology 2021;97(15):e1466– e1474.

14. Forgrave LM, Ma M, Best JR, DeMarco ML. The diagnostic performance of neurofilament light chain in CSF and blood for Alzheimer’s disease, frontotemporal dementia, and amyotrophic lateral sclerosis: A systematic review and meta-analysis. Alzheimer’s & Dementia: Diagnosis, Assessment & Disease Monitoring 2019;11:730–743.

15. Brooks BR, Miller RG, Swash M, Munsat TL. El Escorial revisited: revised criteria for the diagnosis of amyotrophic lateral sclerosis. Amyotrophic lateral sclerosis and other motor neuron disorders : official publication of the World Federation of Neurology, Research Group on Motor Neuron Diseases 2000;1(5):293–299.

16. McKhann GM, Knopman DS, Chertkow H, et al. The diagnosis of dementia due to Alzheimer’s disease: Recommendations from the National Institute on Aging-Alzheimer’s Association workgroups on diagnostic guidelines for Alzheimer’s disease. Alzheimer’s & dementia: the journal of the Alzheimer’s Association 2011;7(3):263–269.

17. Shaw LM, Vanderstichele H, Knapik-Czajka M, et al. Cerebrospinal fluid biomarker signature in alzheimer’s disease neuroimaging initiative subjects [Internet]. Annals of Neurology 2009;65(4):403–413.Available from: https://arxiv.org/abs/PMCID: PMC2696350

18. Crum RM, Anthony JC, Bassett SS, Folstein MF. Population-based norms for the Mini-Mental State Examination by age and educational level. Jama 1993;269(18):2386–2391.

19. Brooks BR. El Escorial World Federation of Neurology criteria for the diagnosis of amyotrophic lateral sclerosis. Subcommittee on Motor Neuron Diseases/Amyotrophic Lateral Sclerosis of the World Federation of Neurology Research Group on Neuromuscular Diseases and th. Journal of the neurological sciences 1994;124:96–107.

20. Montine TJ, Phelps CH, Beach TG, et al. National institute on aging-Alzheimer’s association guidelines for the neuropathologic assessment of Alzheimer’s disease: A practical approach. Acta Neuropathologica 2012;123(1):1– 11.

21. Braak H, Braak E. Neuropathological stageing of Alzheimer-related changes. Acta neuropathologica 1991;82(4):239–259.

22. Brettschneider J, Del Tredici K, Irwin DJ, et al. Sequential distribution of pTDP-43 pathology in behavioral variant frontotemporal dementia (bvFTD). Acta neuropathologica 2014;127(3):423–439.

23. Shaw LM, Vanderstichele H, Knapik-Czajka M, et al. Qualification of the analytical and clinical performance of CSF biomarker analyses in ADNI. Acta neuropathologica 2011;121(5):597–609.

24. Czaplinski A, Yen AA, Appel SH. Forced vital capacity (FVC) as an indicator of survival and disease progression in an ALS clinic population. Journal of Neurology, Neurosurgery & Psychiatry 2006;77(3):390–392.

25. Cedarbaum JM, Stambler N, Malta E, et al. The ALSFRS-R: A revised ALS functional rating scale that incorporates assessments of respiratory function. Journal of the Neurological Sciences 1999;169(1-2):13–21.

26. Quinn C, Edmundson C, Dahodwala N, Elman L. Reliable and efficient scale to assess upper motor neuron disease burden in amyotrophic lateral sclerosis. Muscle & nerve 2020;61(4):508–511.

27. Gromicho M, Figueiral M, Uysal H, et al. Spreading in ALS: The relative impact of upper and lower motor neuron involvement. Annals of clinical and translational neurology 2020;7(7):1181–1192.

28. Visser J, Berg-Vos RM van den, Franssen H, et al. Disease course and prognostic factors of progressive muscular atrophy. Archives of neurology 2007;64(4):522–528.

29. Gordon PH, Cheng B, Katz IB, et al. The natural history of primary lateral sclerosis. Neurology 2006;66(5):647–653.

30. Ahmed RM, Devenney EM, Strikwerda-Brown C, et al. Phenotypic variability in ALS-FTD and effect on survival. Neurology 2020;94(19):e2005–e2013.

31. Karam C, Scelsa SN, Macgowan DJL. The clinical course of progressive bulbar palsy. Amyotrophic Lateral Sclerosis 2010;11(4):364–368.

32. Funder DC, Ozer DJ. Evaluating Effect Size in Psychological Research: Sense and Nonsense [Internet]. Advances in Methods and Practices in Psychological Science 2019;2(2):156–168.Available from: https://doi.org/10.1177/2515245919847202

33. Thiele C, Hirschfeld G. Cutpointr: Improved estimation and validation of optimal cutpoints in R. Journal of Statistical Software 2021;98(11):1–27.

34. Ben-Shachar M, Lüdecke D, Makowski D. effectsize: Estimation of Effect Size Indices and Standardized Parameters [Internet]. Journal of Open Source Software 2020;5(56):2815.Available from: https://github.com/easystats/effectsize

35. Therneau TM. A package for survival analysis in R [Internet]. R package version 2021;Available from: https://cran.r-project.org/package=survival%3E

36. Strong MJ, Yang W, Strong WL, et al. Tau protein hyperphosphorylation in sporadic ALS with cognitive impairment. Neurology 2006;66(11):1770–1771.

37. Strong MJ, Donison NS, Volkening K. Alterations in Tau Metabolism in ALS and ALS-FTSD. Frontiers in Neurology 2020;11:1548.

38. Yang W, Strong MJ. Widespread neuronal and glial hyperphosphorylated tau deposition in ALS with cognitive impairment. Amyotrophic Lateral Sclerosis 2012;13(2):178–193.

39. Conover WJ, Iman RL. Rank transformations as a bridge between parametric and nonparametric statistics. The American Statistician 1981;35(3):124–129.

40. Garg N, Park SB, Vucic S, et al. Differentiating lower motor neuron syndromes. Journal of Neurology, Neurosurgery & Psychiatry 2017;88(6):474–483.

41. Rösler KM, Truffert A, Hess CW, Magistris MR. Quantification of upper motor neuron loss in amyotrophic lateral sclerosis. Clinical neurophysiology 2000;111(12):2208–2218.

42. Cousins KAQ, Phillips JS, Irwin DJ, et al. ATN incorporating cerebrospinal fluid neurofilament light chain detects frontotemporal lobar degeneration. Alzheimer’s & Dementia 2021;17(5):822–830.

43. Gaiani A, Martinelli I, Bello L, et al. Diagnostic and prognostic biomarkers in amyotrophic lateral sclerosis: neurofilament light chain levels in definite subtypes of disease. JAMA neurology 2017;74(5):525–532.

44. Huang F, Zhu Y, Hsiao-Nakamoto J, et al. Longitudinal biomarkers in amyotrophic lateral sclerosis. Annals of clinical and translational neurology 2020;7(7):1103–1116.

45. Illán-Gala I, Lleo A, Karydas A, et al. Plasma tau and neurofilament light in frontotemporal lobar degeneration and Alzheimer disease. Neurology 2021;96(5):e671–e683.

46. Gille B, De Schaepdryver M, Goossens J, et al. Serum neurofilament light chain levels as a marker of upper motor neuron degeneration in patients with amyotrophic lateral sclerosis. Neuropathology and applied neurobiology 2019;45(3):291–304.

47. Al-Chalabi A, Jones A, Troakes C, et al. The genetics and neuropathology of amyotrophic lateral sclerosis. Acta neuropathologica 2012;124(3):339–352.

48. Stevens CH, Guthrie NJ, Roijen M van, et al. Increased tau phosphorylation in motor neurons from clinically pure sporadic amyotrophic lateral sclerosis patients. Journal of Neuropathology & Experimental Neurology 2019;78(7):605–614.

49. Vintilescu CR, Afreen S, Rubino AE, Ferreira A. The Neurotoxic Tau 45-230 Fragment Accumulates in Upper and Lower Motor Neurons in Amyotrophic Lateral Sclerosis Subjects. Molecular Medicine 2016;22(1):477–486.

50. Ashton NJ, Janelidze S, Al Khleifat A, et al. A multicentre validation study of the diagnostic value of plasma neurofilament light. Nature communications 2021;12(1):1–12.

51. Vucic S, Rutkove SB. Neurophysiological biomarkers in amyotrophic lateral sclerosis. Current opinion in neurology 2018;31(5):640–647.

